# COVID-19 convalescent plasma to treat hospitalised COVID-19 patients with or without underlying immunodeficiency: a randomized trial

**DOI:** 10.1101/2022.08.09.22278329

**Authors:** Karine Lacombe, Thomas Hueso, Raphael Porcher, Arsène Mekinian, Thibault Chiarabini, Sophie Georgin-Lavialle, Florence Ader, Julien Saison, Guillaume Martin Blondel, Nathalie De Castro, Fabrice Bonnet, Charles Cazanave, Anne François, Pascal Morel, Olivier Hermine, Valérie Pourcher, Marc Michel, Xavier Lescure, Nora Soussi, Philippe Brun, Fanny Pommeret, Pierre-Olivier Sellier, Stella Rousset, Lionel Piroth, Jean-Marie Michot, Gabriel Baron, Xavier De Lamballerie, Xavier Mariette, Pierre-Louis Tharaux, Matthieu Resche-Rigon, Philippe Ravaud, Tabassome Simon, Pierre Tiberghien

## Abstract

**Objectives:** Efficacy of convalescent plasma in COVID-19 pneumonia (CPP) is uncertain, especially in immunocompromised patients. CORIMUNO-CORIPLASM is an open-label, Bayesian randomised clinical trial embedded in the CORIMUNO trials platform that evaluated the efficacy of CCP in patients with moderate COVID-19.

**Setting:** 19 university and general hospitals across France.

**Participants:** Adult hospitalized with a positive SARS-CoV2 test, duration of symptoms < 9 days and WHO score severity 4 or 5 who signed written inform consent.

**Intervention:** Open label randomisation to either usual care (UC) or 4 units (200-220 ml/unit, 2 units/day over 2 consecutive days) of convalescent plasma (CCP) with a seroneutralisation titer > 40.

**Outcomes:** Primary outcome was proportion of patients with a WHO-Clinical Progression Score (CPS) ≥6 on the 10-point scale on day (d) 4 (higher values indicating a worse outcome) and survival without ventilation or additional immunomodulatory treatment by day 14. Secondary outcomes included evolution of WHO-CPS, overall survival, time to discharge and time to oxygen supply independency. Pre-defined subgroups analyses included immunosuppression status, duration of symptoms before randomization and use of steroids.

**Results:** A total of 120 patients were recruited and assigned to CCP (n=60) or UC (n=60), including 22 (CCP) and 27 (UC) immunocompromised patients. Thirteen (22%) patients with CCP had a WHO-CPS ≥6 at day 4 versus 8 (13%) with UC, adjusted odds ratio (aOR) 1.88 [95%CrI 0.71 to 5.24]. By day 14, 19 (31.6%) patients with CCP and 20 (33.3%) patients with UC had ventilation, additional immunomodulatory treatment or had died. Cumulative incidence of death was 3 (5%) with CCP and 8 (13%) with UC at day 14 (aHR 0.40 [95%CrI 0·10 -1·53]), and 7 (12%) with CCP and 12 (20%) with UC at day 28 (aHR 0.51 [95%CrI 0.20-1.32]). I n a s ubgroup analysis performed in immunocompromised patients, the association of CCP with mortality was HR 0.39 [95%CI 0.14-1.10].

**Conclusions:** CCP did not improve early outcomes in patients with moderate COVID-19. Its efficacy in immunocompromised patients needs to be further explored.

**Trial registration:** clinicaltrials.gov Identifier: NCT04345991

**KEY MESSAGES BOX:** *What is already known on this topic?:*

- Convalescent plasma treatment, i.e., passive polyclonal antibody administration to provide immediate immunity, has been used to improve the survival rate of patients with severe acute respiratory syndromes of viral etiology in emergency settings and times where there was no specific antiviral treatment
- At the early stages of the COVID-19 pandemic, using high titre COVID-19 convalescent plasma (CCP) appeared to be an immediate therapeutic option.
- However, a large number of randomised clinical trials and observational studies have yielded conflicting results regarding the efficacy of CCP.
- Furthermore, the efficacy of CCP in patients with underlying immunosuppression has been evaluated only in a limited manner.
- The emergence of variants resistant to other passive immunotherapy approaches, ie monoclonal antibodies, has limited the therapeutics options for such patients

*What this study adds ?:*

- This multicentre randomised clinical trial provided evidence that high titre CCP in a population hospitalised with a mild to moderate form of COVID-19 within 9 days of symptoms onset may not improve early outcome.
- In the subgroup of patients with immunosuppression, there was evidence suggesting a lower odds of death 14 and 28 days after CCP transfusion, albeit without reaching significance.

*How does this study might affect research, practice of policy:*

- The result of study, along with the recent data obtained from other trials and cohort studies supports further evaluation of CCP transfusion in patients with underlying immunosuppression for whom therapeutic options are currently scarce if non-existent, due to the ever changing genetic variability of SARS-CoV2.

## INTRODUCTION

Early in the COVID-19 pandemic, COVID-19 convalescent plasma (CCP) transfusion was identified as a potential treatment that needed evaluation. (1) Overall efficacy of CCP in hospitalized patients has not been established.(2) However, high titre CCP may be beneficial particularly if used early before seroconversion (3,4) or in patients unable to mount an effective humoral response.(5,6) Monoclonal antibody treatment has demonstrated efficacy as an early intervention (7) or later in hospitalised seronegative patients,(8) however with significant limitations including accessibility and cost,(9) as well as loss of efficacy as recently exemplified with the emergence of the immune – evading omicron SARS-CoV-2 subvariants.(10) By contrast, CCP from convalescent vaccinated donors is cheaper, readily available and adaptable to a changing viral landscape, and potentially less prone to immune resistance. Indeed, while the recent omicron waves have been associated with a steep decrease in the efficacy of almost all available monoclonal antibodies (11), high titre CCP from (pre-omicron) convalescent vaccinated donors may retain anti-omicron neutralization activity. (12) Such anti-micron neutralisation capacity is further increased in CCP from omicron convalescent vaccinated donors.(13) Alongside immunomodulating drugs that specifically target the inflammatory phase of the disease, oral direct antiviral agents such as molnupiravir (14) or nirmatrelvir/ritonavir (15) also represent another therapeutic option. Such drugs have however drawbacks, such as necessitating an initiation within 5 days of symptoms onset and drug interactions for nirmatrelvir/ritonavir, notably in immunosuppressed patients. Lastly, the intravenous antiviral agent remdesivir has demonstrated only limited anti-SARS-CoV-2 efficacy in hospitalised patients.(16) Careful assessment of CCP efficacy and safety therefore remains an important public health issue, particularly in immunosuppressed patients unable to mount a vaccine-mediated immune response and at risk of severe disease with limited therapeutic options. We report the results of a randomized controlled trial assessing the efficacy of CCP (4 units, ≈ 840 ml) in immune-competent and immunosuppressed patients hospitalized for moderate SARS-CoV-2 associated pneumonia requiring no assisted ventilation at time of inclusion.

## METHODS

### Trial design and study oversight

CORIMUNO-19 is a platform trial established by Assistance Publique-Hôpitaux de Paris, France, at the early beginning of the COVID-19 pandemic.(17) CORIMUNO-CORIPLASM was an embedded multicentric, open-label randomized controlled trial in patients with moderate COVID-19 pneumonia conducted in French hospitals across France (NCT04345991). Ethical clearance was obtained from CPP Ile de France VI on April 10, 2020 (no. 26-20 Med.1°). The full trial protocol and statistical analysis plan is available in Appendix II and III.

### Study population and randomization

At hospital admission, patients were evaluated for eligibility criteria: hospitalized adult ≥18 years of age, positive SARS-CoV-2 nasopharyngeal PCR and/or CT scan prior to randomisation, onset of symptoms <9 days, illness of mild or moderate severity according to the WHO clinical progression scale (CPS) (hospitalised, mild disease: no oxygen need; hospitalised, moderate disease: oxygen need <3l; (Appendix I), no pregnancy, no prior severe grade 3 allergic reaction to plasma transfusion, and no current documented bacterial infection. ABO compatibility with available CCP was verified before patient inclusion. Written informed consent was obtained from all patients or their legal representatives at inclusion in CORIMUNO19. A specific written informed consent was sought from eligible patients before inclusion in the CORIPLASM trial. The independent clinical research organisation drew up the computerized randomization list, and the patient’s randomization number was accessed through a secure site by a site study team member. Randomisation was performed within 2 hours after enrolment. Eligible patients were randomised 1:1 to receive either convalescent plasma or usual care. The latter could include the use of dexamethasone, tocilizumab, supportive care including supplemental oxygen, antivirals, and antibiotics. A Data and Safety Monitoring Board (DSMB) provided guidance to the trial after inclusion of every 60 patients.

### Study product

Convalescent donors were eligible for plasma donation 15 days after the resolution of COVID-19-related symptoms. Collected apheresis plasma by Etablissement Français du Sang (EFS) underwent pathogen reduction (INTERCEPT Blood System, Cerus, Concord, CA) and standard testing as per current regulations in France. Anti-SARS-CoV-2 potency was assessed in each donation, with a requirement for a SARS-CoV-2 seroneutralization titer ≥=40, as described in (18). Additionally, antibody content was determined by IgG enzyme-linked immunosorbent assay (ELISA EUROIMMUN, Bussy-Saint-Martin, France). CCP with a seroneutralization titer >=40 and made available for the trial, all collected between April and June 2020, yielded a mean ELISA ratio of 6.1 (s.d.: 2.9, IQR: IQR: 5.4, min. 0.4, max: 13.0). After the first 3 patients received 2 units of ABO-compatible CCP as per protocol, all subsequent patients randomised to the CCP arm received 4 units of CCP (200-220 ml/unit, 2 units/day over 2 consecutive days) provided by different donors.

### Study endpoints

As in all CORIMUNO19 nested trials, there was an early primary endpoint defined as a WHO Clinical Progression Scale (WHO-CPS) score ≥ 6 (Supp. material, Appendix I) at day 4 of randomisation, higher values of the WHO-CPS indicating a worse outcome. The primary endpoint specific to the CORIPLASM trial was survival without the need for ventilator use (including non-invasive ventilation, NIV or high flow oxygen) at day 14 of randomization (WHO-CPS < 6) or additional immunomodulatory treatment, with the exception of corticosteroids included within the standard of care (study amendment, appendix). Secondary endpoints included WHO-CPS at 4, 7 and 14 days after randomization, overall survival at 14 and 28 days after randomisation (i.e. for the periods from day 1 to day 14 and from day 1 to day 28, respectively), time to discharge, time to oxygen supply independency and evolution of a series of biological parameters at days 4, 7 and 14 after randomization. Pre-defined subgroups analyses included immunosuppression status (underlying immunodeficiency: yes/no), duration of symptoms before randomization (≤5 days, > 5 days), and use of steroids. Safety data included all clinical and biological adverse events observed during study follow-up. Immunodeficiency was defined as the presence of at least one of the following medical conditions: active malignant neoplasm, lymphoid or myeloid neoplasms, hematopoietic stem cell or solid organ transplantation, or HIV/AIDS not on highly active antiretroviral treatment.

### Statistical Analysis

The sample size was set at 120 participants (60 per group), with a Bayesian interim analysis after 60 participants were randomised. We computed that the trial would have a frequentist power of 97.2% to detect a decrease in event proportion from 0.50 to 0.20, and 73.9% to detect a decrease in event proportions from 0.50 to 0.30. The study statisticians, who were masked to the group assignment, oversaw the interim and final analyses. Interim analysis reports were only shared with DSMB members and not with trial investigators, who remained blinded to all results during the trial.

The treatment effect was primarily expressed as an absolute risk difference (ARD) for the early primary endpoint, and a hazard ratio (HR) for the longer-term primary endpoint. Both were analysed in a Bayesian framework. A posterior probability of ARD <0 or HR <1 greater than 0.99 at the interim analysis or greater than 0.95 at the final analysis indicated efficacy. We also computed posterior probabilities of ARD <−5.5% and HR <0.85, denoting a moderate or greater effect. At the interim analysis, a posterior probability of moderate or greater impact <0.20 defined a futility boundary. The treatment effect was summarised by the posterior median and equal tail credible intervals (CrIs). Because the decision rules are one-sided, consistent CrIs would theoretically be one-sided 95% CrIs, but we chose to report two-sided 90% CrIs with the same upper bound. For the early primary endpoint, the posterior distribution of ARD was computed analytically, using a beta prior distribution with parameters 1 and 1 for the proportion in each group. An odds ratio (OR) adjusted for age and centre, the latter being treated as a random effect. was also estimated using a Bayesian logistic regression model. For the longer-term primary endpoint, the posterior HR distribution adjusted for age and centre was computed using Markov chain Monte Carlo (MCMC) with Gaussian prior distributions with mean 0 and variance 10^6^ for the log HR. Different prior distributions were used as sensitivity analyses. Secondary outcomes were analysed in a frequentist framework, except for WHO-CPS scores, analysed as an ordinal variable with a Bayesian proportional odds model. Analyses of secondary outcomes were also adjusted for age and centre. For time to discharge and time to oxygen supply independency, we estimated adjusted sub distribution hazard ratios (SHRs) using Fine-Gray models, death being the competing event. Estimating SHRs was preferred over cause-specific HRs because they have a one-to-one relationship with the cumulative incidence, i.e. the proportion, of events, and we considered they would therefore be more relevant that the ratio of rates at which those events occur in time. Interaction tests between the treatment group and subgroups were used to tests for treatment effect heterogeneity between subgroups, using similar regression models as the ones for the main adjusted analyses. Details on the statistical analyses are given in the Statistical Analysis Plan.

Analyses were done on an intention-to-treat (ITT) basis. The original protocol specified a modified ITT analysis excluding patients declining the intervention and those unable to receive planned plasma therapy due to unavailability of ABO-compatible CCP. Since those situations did not occur, no modified ITT analysis was performed. No correction for multiplicity was done for secondary outcomes, and corresponding results should be regarded as exploratory.

Two interim analyses were conducted (Table S1). Statistical analyses used SAS (version 9.4, SAS Institute) and R (version 4.0.5, R Foundation) statistical software.

## RESULTS

### Study population and CCP administration

Between April 16, 2020, and April 21, 2021, a total of 120 patients (60 with CCP, 60 with UC only) were enrolled (Figure S1). Subjects’ characteristics are reported in Table 1 appear well balanced between both groups.

**Table 1.**
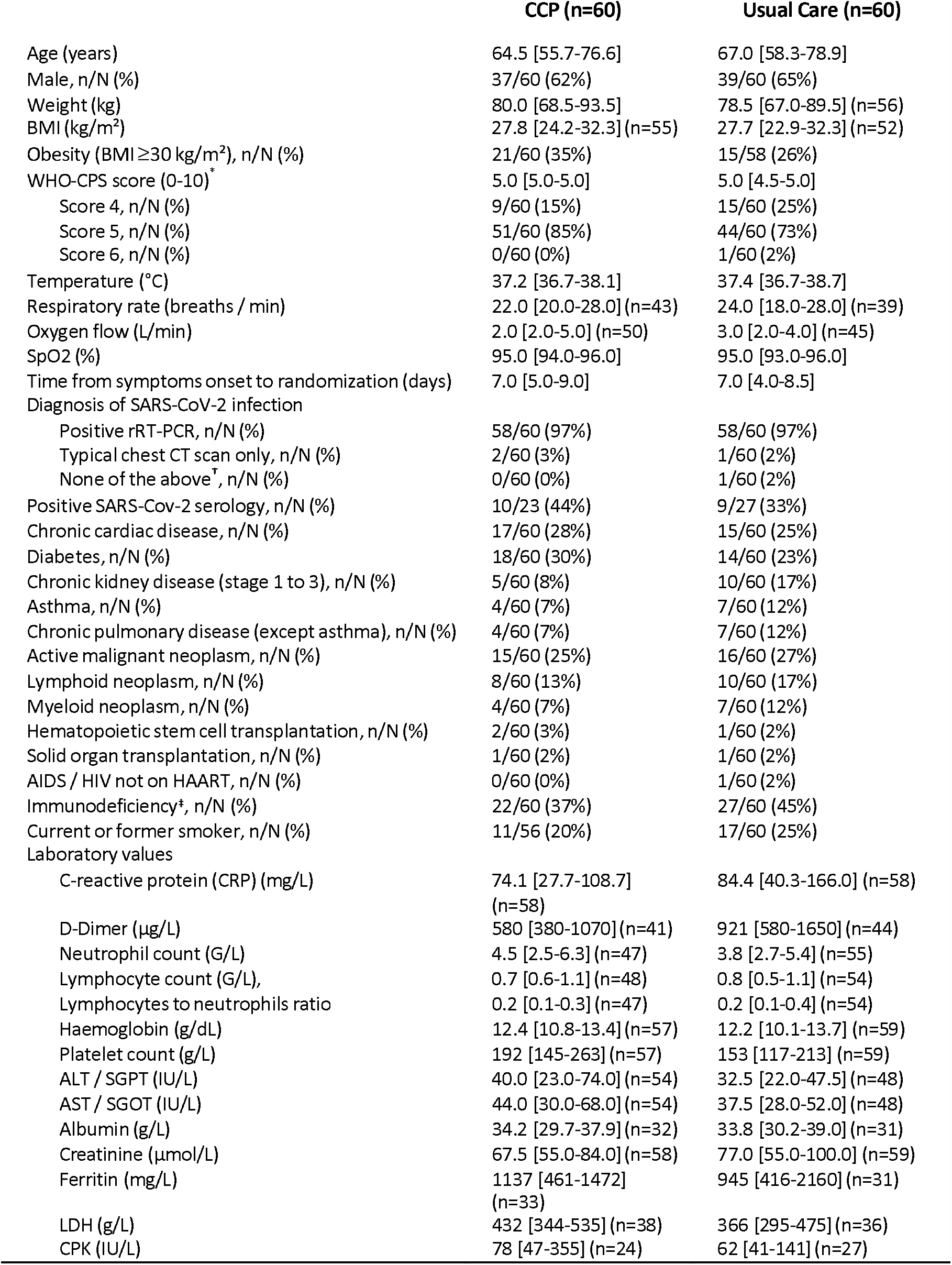
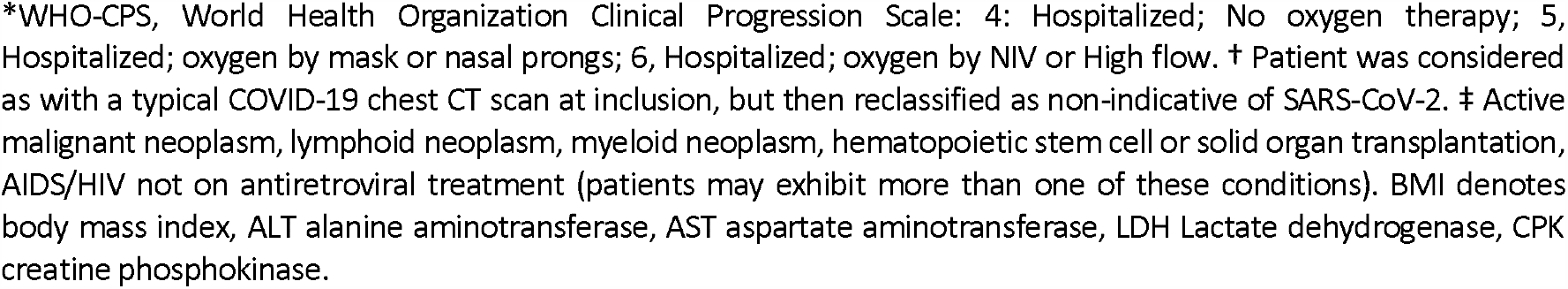
Characteristics at baseline. Values are median [interquartile range] unless stated otherwise.

|  | CCP (n=60) | Usual Care (n=60) |
| --- | --- | --- |
| Age (years) | 64.5 [55.7-76.6] | 67.0 [58.3-78.9] |
| Male, n/N (%) | 37/60 (62%) | 39/60 (65%) |
| Weight (kg) | 80.0 [68.5-93.5] | 78.5 [67.0-89.5] (n=56) |
| BMI (kg/m<sup>2</sup>) | 27.8 [24.2-32.3] (n=55) | 27.7 [22.9-32.3] (n=52) |
| Obesity (BMI ≥30 kg/m<sup>2</sup>), n/N (%) | 21/60 (35%) | 15/58 (26%) |
| WHO-CPS score (0-10)<sup>*</sup> | 5.0 [5.0-5.0] | 5.0 [4.5-5.0] |
| Score 4, n/N (%) | 9/60 (15%) | 15/60 (25%) |
| Score 5, n/N (%) | 51/60 (85%) | 44/60 (73%) |
| Score 6, n/N (%) | 0/60 (0%) | 1/60 (2%) |
| Temperature (°C) | 37.2 [36.7-38.1] | 37.4 [36.7-38.7] |
| Respiratory rate (breaths / min) | 22.0 [20.0-28.0] (n=43) | 24.0 [18.0-28.0] (n=39) |
| Oxygen flow (L/min) | 2.0 [2.0-5.0] (n=50) | 3.0 [2.0-4.0] (n=45) |
| SpO<sub>2</sub> (%) | 95.0 [94.0-96.0] | 95.0 [93.0-96.0] |
| Time from symptoms onset to randomization (days) | 7.0 [5.0-9.0] | 7.0 [4.0-8.5] |
| Diagnosis of SARS-CoV-2 infection |  |  |
| Positive rRT-PCR, n/N (%) | 58/60 (97%) | 58/60 (97%) |
| Typical chest CT scan only, n/N (%) | 2/60 (3%) | 1/60 (2%) |
| None of the above<sup>†</sup>, n/N (%) | 0/60 (0%) | 1/60 (2%) |
| Positive SARS-Cov-2 serology, n/N (%) | 10/23 (44%) | 9/27 (33%) |
| Chronic cardiac disease, n/N (%) | 17/60 (28%) | 15/60 (25%) |
| Diabetes, n/N (%) | 18/60 (30%) | 14/60 (23%) |
| Chronic kidney disease (stage 1 to 3), n/N (%) | 5/60 (8%) | 10/60 (17%) |
| Asthma, n/N (%) | 4/60 (7%) | 7/60 (12%) |
| Chronic pulmonary disease (except asthma), n/N (%) | 4/60 (7%) | 7/60 (12%) |
| Active malignant neoplasm, n/N (%) | 15/60 (25%) | 16/60 (27%) |
| Lymphoid neoplasm, n/N (%) | 8/60 (13%) | 10/60 (17%) |
| Myeloid neoplasm, n/N (%) | 4/60 (7%) | 7/60 (12%) |
| Hematopoietic stem cell transplantation, n/N (%) | 2/60 (3%) | 1/60 (2%) |
| Solid organ transplantation, n/N (%) | 1/60 (2%) | 1/60 (2%) |
| AIDS/ HIV not on HAART, n/N (%) | 0/60 (0%) | 1/60 (2%) |
| Immunodeficiency<sup>‡</sup>, n/N (%) | 22/60 (37%) | 27/60 (45%) |
| Current or former smoker, n/N (%) | 11/56 (20%) | 17/60 (25%) |
| Laboratory values |  |  |
| C-reactive protein (CRP) (mg/L) | 74.1 [27.7-108.7] (n=58) | 84.4 [40.3-166.0] (n=58) |
| D-Dimer (µg/L) | 580 [380-1070] (n=41) | 921 [580-1650] (n=44) |
| Neutrophil count (G/L) | 4.5 [2.5-6.3] (n=47) | 3.8 [2.7-5.4] (n=55) |
| Lymphocyte count (G/L), | 0.7 [0.6-1.1] (n=48) | 0.8 [0.5-1.1] (n=54) |
| Lymphocytes to neutrophils ratio | 0.2 [0.1-0.3] (n=47) | 0.2 [0.1-0.4] (n=54) |
| Haemoglobin (g/dL) | 12.4 [10.8-13.4] (n=57) | 12.2 [10.1-13.7] (n=59) |
| Platelet count (g/L) | 192 [145-263] (n=57) | 153 [117-213] (n=59) |
| ALT / SGPT (IU/L) | 40.0 [23.0-74.0] (n=54) | 32.5 [22.0-47.5] (n=48) |
| AST / SGOT (IU/L) | 44.0 [30.0-68.0] (n=54) | 37.5 [28.0-52.0] (n=48) |
| Albumin (g/L) | 34.2 [29.7-37.9] (n=32) | 33.8 [30.2-39.0] (n=31) |
| Creatinine (µmol/L) | 67.5 [55.0-84.0] (n=58) | 77.0 [55.0-100.0] (n=59) |
| Ferritin (mg/L) | 1137 [461-1472] (n=33) | 945 [416-2160] (n=31) |
| LDH (g/L) | 432 [344-535] (n=38) | 366 [295-475] (n=36) |
| CPK (IU/L) | 78 [47-355] (n=24) | 62 [41-141] (n=27) |

\*WHO-CPS, World Health Organization Clinical Progression Scale: 4: Hospitalized; No oxygen therapy; 5, Hospitalized; oxygen by mask or nasal prongs; 6, Hospitalized; oxygen by NIV or High flow. † Patient was considered as with a typical COVID-19 chest CT scan at inclusion, but then reclassified as non-indicative of SARS-CoV-2. ‡ Active malignant neoplasm, lymphoid neoplasm, myeloid neoplasm, hematopoietic stem cell or solid organ transplantation, AIDS/HIV not on antiretroviral treatment (patients may exhibit more than one of these conditions). BMI denotes body mass index, ALT alanine aminotransferase, AST aspartate aminotransferase, LDH Lactate dehydrogenase, CPK creatine phosphokinase.

The median time between the onset of symptoms and CCP transfusion was 7 days in both groups. A positive anti-S and anti-N SARS-CoV-2 serology was noted in 10/23 (44%) evaluable patients receiving CCP and 9/27 (33%) evaluable patients receiving UC. Twenty two /sixty (45%) and 27/60 (37%) patients had an underlying immunodeficiency in the CCP and UC arms, respectively. One patient was considered to have COVID-19 with a typical chest CT scan at inclusion, but was then reclassified as non-indicative of SARS-CoV-2 infection and finally diagnosed with pulmonary oedema from cardiac origin. The other treatments received before and after randomisation until day 14 are reported in Table S2. Intention to treat analysis was performed on 120 patients, of whom 2 in each study group were lost to follow-up at day 28 evaluation but discharged alive before day 28 (Figure 1). One patient did not receive any plasma infusion because of sudden worsening after randomisation and transfer to an intensive care unit (ICU), 9 patients received 2 units (3 as per protocol, 6 because of worsening of clinical status leading to ICU admission), and 50 received 4 units. Same day transfusion occurred in 78% of patients, whereas 12 (20%) and 1 (2%) were transfused 1 and 3 days after randomisation, respectively.

**Figure 1:**
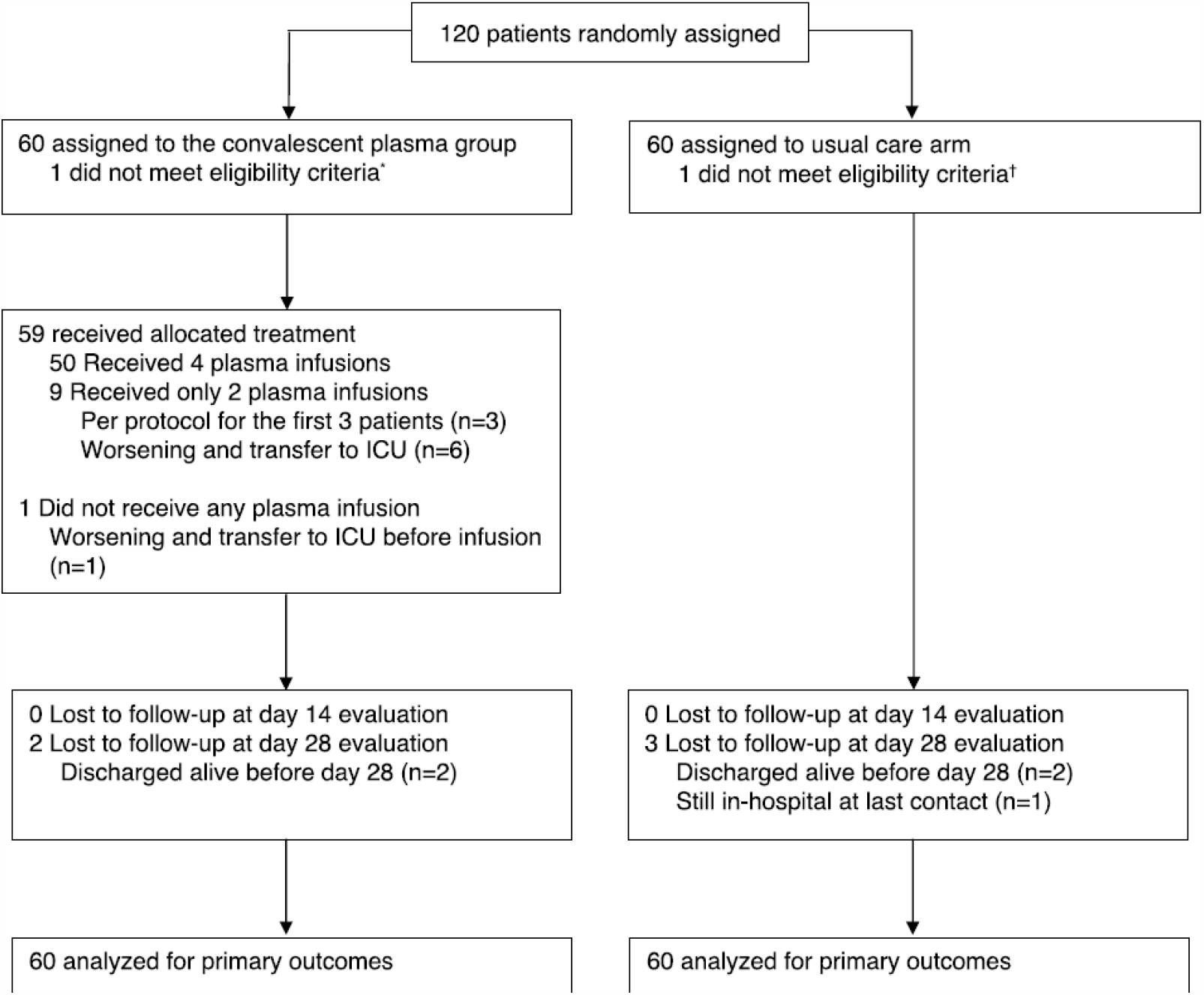
Flow chart of the CORIPLASM clinical trial. * No SARS-CoV-2 infection; † Under non-invasive ventilation.

### Primary outcomes

Thirteen (22 %) patients in the CCP arm versus 8 (13%) patients in the UC arm had a WHO-CPS ≥6 at day 4 (median posterior absolute risk difference +8·0%; 90% credible interval [CrI] –3.2 to +19.4), adjusted odds ratio (aOR) 1.88 (95% CrI 0.71–5.24) (Tables 2 and S3, Figure S2). Analysis of the day 4 WHO-CPS score was analysed as an ordinal outcome in a proportional odds model, yielded a median posterior adjusted odds ratio of 1·42 (95% CrI 0·70–2·91), therefore showing higher scores in the CCP arm, although the difference was not significant. By day 14, 19 (31.6%) and 20 (33.3%) patients in the CCP and UC arms, respectively, needed non-invasive or high flow ventilation (CCP:15, UC:13) or additional immunomodulatory treatment (anti-IL-6 receptor monoclonal antibody) (CCP:0, UC:5) or had died (2 in each arm, in addition to 1 and 5 deaths that occurred after reaching the primary outcome in the CCP and UC arm, respectively). The cumulative incidence of ventilation or death is reported in Figure 2a. The median posterior adjusted hazard ratio was 1.04 [95% CrI 0.55-1.97], and the posterior probability of a moderate or greater benefit was 0.269 (Tables 2 and S4). Results remained highly consistent across the range of prior distributions used in sensitivity analyses (figure S3).

**Table 2.** Primary and secondary efficacy outcomes.

|  | Convalescent plasma (n=60) | Usual care (n=60) | Treatment effect |
| --- | --- | --- | --- |
| **Co-primary outcomes** |  |  |  |
| WHO-CPS score $\geq 6$ at day 4 | 13 (22%) | 8 (13%) | +8.0% (90% CrI -3.2 to +19.4)* |
| Posterior probability of any benefit |  |  | 11.9% |
| Posterior probability of moderate or greater benefit<sup>x</sup> |  |  | 2.4% |
| Need for ventilation, additional immunomodulators or death up to day 14 | 19 (32%) | 20 (33%) | 1.04 (90% CrI 0.61 to 1.78)<sup>†</sup> |
| Posterior probability of any benefit |  |  | 45.2% |
| Posterior probability of moderate or greater benefit<sup>x</sup> |  |  | 26.9% |
| **Secondary outcomes** |  |  |  |
| Overall survival |  |  |  |
| Mortality over day 0–14 | 3 (5%) | 8 (13%) | 0.40 (95% CI 0.10 to 1.53)<sup>‡</sup> |
| Mortality over day 0–28 | 7 (12%) | 12 (20%) | 0.51 (95% CI 0.20 to 1.32)<sup>‡</sup> |
| WHO-CPS score (10 pt-scale) |  |  |  |
| Day 4, median [IQR] | 5 [5–5] | 5 [4–5] | 1.42 (95% CrI 0.70 to 2.91)<sup>¶¶</sup> |
| Day 7, median [IQR] | 5 [4–5]** | 5 [4–5]<sup>††</sup> | 1.20 (95% CrI 0.61 to 2.37)<sup>¶¶</sup> |
| Day 14, median [IQR] | 3 [2–4]<sup>‡‡</sup> | 3 [2–5]<sup>††</sup> | 0.59 (95% CrI 0.30 to 1.13)<sup>¶¶</sup> |
| Day 2 to 14 (longitudinal analysis) | — | — | 1.04 (95% CrI 0.37 to 2.86)<sup>¶¶</sup> |
| Time to discharge |  |  |  |
| Discharged at day 28 | 48 (80%) | 45 (75%) | 0.99 (95% CI 0.65 to 1.49)<sup>¶¶¶</sup> |
| Time to oxygen supply independency<sup>x</sup> |  |  |  |
| Independent from oxygen at day 28 | 42/51 (82%) | 32/45 (71%) | 1.18 (95% CI 0.73 to 1.91)<sup>¶¶¶</sup> |

CrI: credible interval (Bayesian analysis); CI: confidence interval (frequentist analysis); WHO-CPS: World Health Organization Clinical Progression Scale. Moderate or greater benefit was defined as an absolute risk difference $< -5.5\%$ for the d4 outcome and a hazard ratio $< 0.85$ for the d14 outcome. \* Median posterior absolute risk difference; the median posterior odds ratio adjusted for age and centre was 1.88 (90% CrI 0.83 to 4.44).
<sup>†</sup> Median posterior hazard ratio adjusted for age and centre. <sup>‡</sup> Hazard ratio adjusted for age and centre.
<sup>¶¶</sup> Median posterior odds ratio in a proportional odds model adjusted for age and centre. \*\* n=58 with available data. <sup>††</sup> n=59 with available data. <sup>‡‡</sup> n=59 with available data. <sup>¶¶¶</sup> Subdistribution hazard ratio adjusted for age and centre. <sup>x</sup> For participants needing oxygen at randomisation (WHO-CPS $\geq 5$ ).

**Figure 2:**
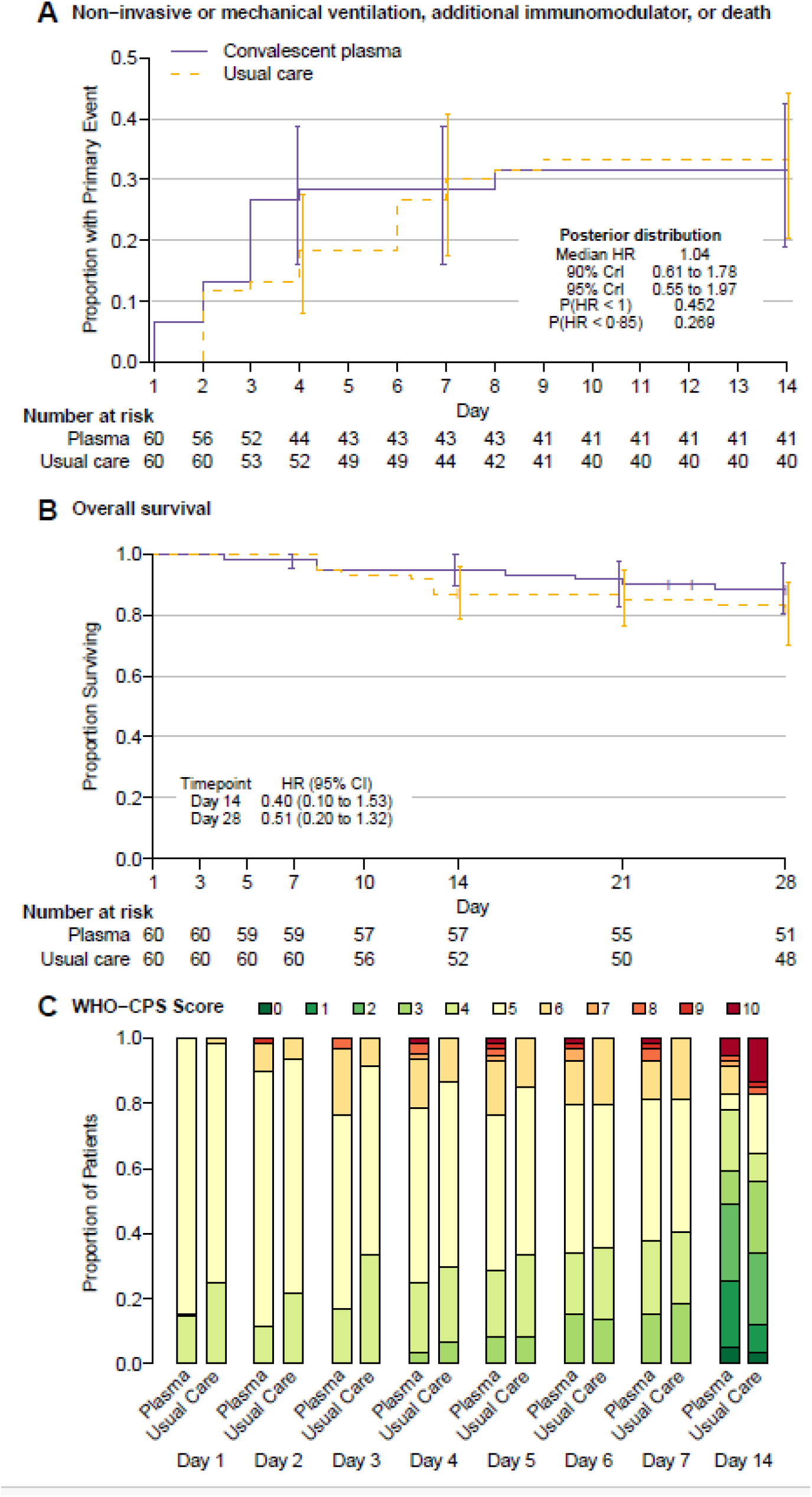
Study outcomes. **2a**. Cumulative incidence of non-invasive or mechanical ventilation or use of additional immunomodulatory drugs or death over 14 days (events at day 1 of randomization occurred on the same day, but after randomisation); **2b**. Overall survival during follow-up; **2c**.Distribution of WHO-CPS during follow-up.

### Secondary outcomes

At day 14, the cumulative incidence of death was 3 (5%) and 8 (13%) in the CCP and UC arms, respectively (aHR 0·40 [95% confidence interval (CI) 0.10-1.53])(Figure 2b, Table S5). At day 28, 7 (12%) and 12 (20%) patients had died in the CCP and UC groups, respectively, with aHR 0.51 [95% CI 0.20-1.32]. The distribution of the WHO-CPS from day 1 to day 14 did not significantly differ within groups, with a posterior odds ratio of 1·04 (95% CrI 0·37– 2·86) for the CCP group compared to UC in a longitudinal ordinal model. More precisely, WHO-CPS scores tended to be higher in the CCP group between day 3 and 5, and then lower at day 14, with a lower mortality in particular (figure 2c, Table S6), albeit not significant. At day 14 and day 28, 38 and 48 patients in the CCP group and 36 and 45 in the UC group were discharged, respectively, with an adjusted day 28 sub distribution hazard ratio (SHR) of 0.99 [95% CI 0.65-1.49] adjusted for age and centre sub-distribution (Table S7). The incidence of oxygen supply independency until day 28 was not different between groups: 76% and 62% by day 14, 82% and 71% by day 28, in the CCP and UC arms, respectively (SHR: 1.18, [95% CI 0.73-1.91])(Table S7).

### Subgroup analyses

Figure 3a reports the primary outcome at day 14, with no difference in the different subgroups. In the 47 patients who had an underlying immunodeficiency, the rate of a WHO-CPS ≥6 at day 4 was not statistically different in the CCP arm compared to the UC arm (24% versus 15%, aOR: 1.97 [95% CI 0.53-7.39]). At day 28, 4 of 21 patients had died in the CCP group versus 9 out of 26 in the UC group (HR 0.39, [95% CI 0.14-1.10])(Figure 3b). Despite these findings favouring CCP, there was no evidence of an interaction between immunodeficiency status and treatment (p=0.34): 4/21 patients had died in the CCP group versus 9/26 in the UC group (HR 0.39, [95% CI 0.14-1.10])(Figure 3b). Limited mortality was observed in the absence of underlying immunodeficiency (Figure 3c). Neither the symptoms duration nor the use of dexamethasone had an impact on day 28 survival (Table S8). Post-hoc analysis of antibody potency in transfused CCP in relation to outcome did not reveal a significant dose-effect (Table S9).

**Figure 3:**
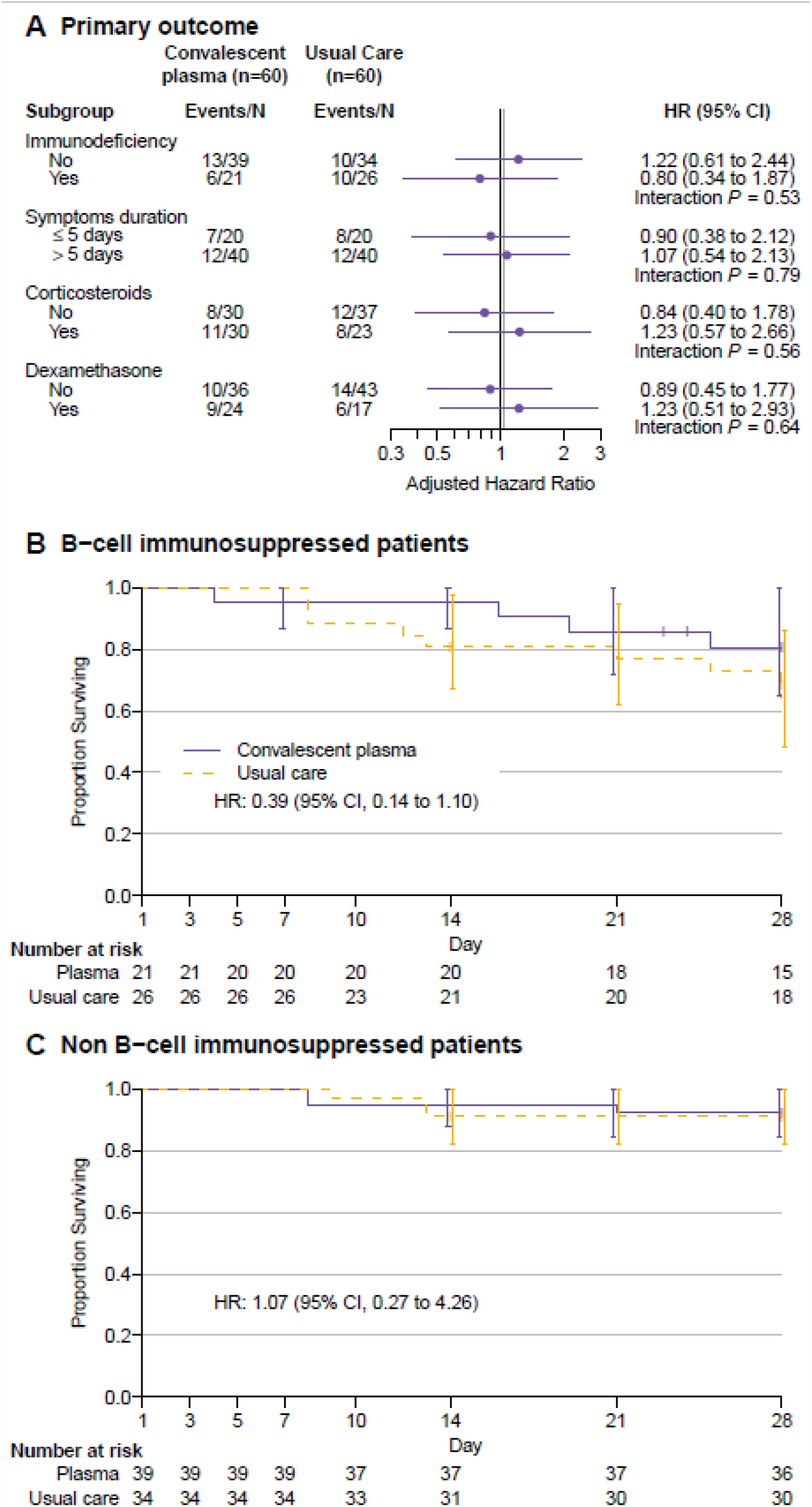
Subgroup analyses. **3a**. Day 14 primary outcomes (need for non-invasive or mechanical ventilation or use of additional immunomodulatory drugs or death). The grey line indicates the overall estimate of treatment effect. Given only one patient was receiving antivirals at randomisation, no subgroup analysis according to antivirals was performed; **3b and 3c**. Overall survival during follow-up in patients with (3b) or without (3c) underlying immunodeficiency.

### Safety

Adverse events were reported in 44 (73%) and 36 (60%) patients in the CCP (n=124) and UC (n=103) arms, respectively: incidence rate ratio = 1.06 [95% CI 0.63-1.77])(Table S9). Serious adverse events were noted in 30 (50%) and 26 (43%) patients in the CCP (n=46) and UC (n=48) arms, respectively (incidence rate ratio = 0.84 [95%CI 0.46-1.54]). Of note, 10 sepsis-related events were observed with UC (6 with CCP) and 4 acute pulmonary oedema were reported with CCP (none with UC). Causes of death were COVID-19-related acute respiratory distress syndrome (UC:10, CCP:3), cardiac origin (UC:0,CCP:2), sepsis (CCP:2, UC:3), gastro-intestinal (CCP:0, UC:1), vascular (CCP:1, UC:0), and 1 of unknown origin (CCP).

## DISCUSSION

In CORIMUNO-CORIPLASM trial, no difference was found in terms of early outcome between CCP and usual care for hospitalised COVID-19 patients not requiring assisted ventilation. The survival rate at day 14 and again at day 28 was numerically higher in the CCP arm, however without reaching statistical significance.

The absence of efficacy associated with CCP agrees with the results of most prospective randomized clinical trials currently reported in hospitalized COVID-19 patients.(19) Indeed, only a limited number of randomized studies have reported a better survival following CCP treatment,(20,21) while several other trials, notably the large RECOVERY trial,(22) have found no evidence of survival benefit with CCP. Reasons for these discrepancies may relate to CCP characteristics, time to treatment from first symptoms, treatment modalities, and patient characteristics. Of note, large retrospective studies in the US reported evidence of reduced mortality associated with CCP treatment in patients hospitalized with COVID-19 (23,24).

Importantly, our study included a significant proportion of patients with underlying immunosuppression. In agreement with prior findings,(25) such COVID-19 patients have a worse prognosis, as noted in the UC arm. Several studies have suggested that CCP may be particularly effective in patients unable to mount an immune response, notably a humoral response. We reported earlier that CCP treatment in immunosuppressed patients, mainly B-cell haematological malignancies treated by anti-CD-20 monoclonal antibodies, was associated with a favourable outcome.(6) Further evidence was provided by two independent exposed/non-exposed studies with propensity score in patients with underlying immunosuppression. Hazard ratios of 0.52 [95%CI 0.29-0.92] and 0.50 [95%CI 0.34-0.72] were reported in favour of CCP treatment, respectively.(6,7) Such reduction in mortality is strikingly close to the reduction in mortality observed in similar patients randomized to the CCP arm of the CORIPLASM trial.

Most of the other randomised trials published to date did not report subgroup analysis for patients with underlying immunosuppression. One notable exception is the REMAP-CAP trial that investigated CCP in critically ill COVID-19 patients.(22) Although the overall results of this study did not provide evidence of the efficacy of CCP in such patients, a pre-specified subgroup analysis revealed a potential benefit of CCP in patients with immunodeficiency. Another randomized trial performed in patients with a diverse range of clinical conditions reported a significant effect of CCP on clinical improvement and survival in the subgroup of 56 individuals with cancer (26). Finally, a meta-analysis incorporating the present trial has confirmed the potential benefit of CCP in immunosuppressed individuals with mild to moderate Covid19, thanks to more statistical power due to the high number of immunosuppressed individuals included (1487 from randomized trials, 265 from case series and 368 from cohorts).(27)

An antibody (Ab)-dose effect has been evidenced in several randomized studies (3,28) as well as in the early access program in the United States.(23) In the CORIPLASM study, 800-880 ml of CCP were transfused to patients randomised to the CCP arm. In most reported studies to date, patients most often received 250 ml-500 ml of CCP, with the notable exception of the CAPSID trial where CCP patients received 700-750 ml.(28)^24^ Interestingly, the CAPSID trial reported a significant Ab-dose effect regarding several outcomes, including survival at day 28. Differently from CAPSID, the CORIPLASM protocol recommended four CCP units provided by different donors for each patient, which resulted in less variation in mean Ab content in transfused CCP from patient to patient. This difference in transfusion practice may have contributed to reducing the ability to identify an Ab dose effect in our study. Furthermore, CCP for the CORIPLASM study were collected early in the COVD-19 crisis, when vaccination was not yet available, and prior to the occurrence of relevant SARS-CoV-2 variants.

Several studies have demonstrated that plasma provided by convalescent vaccinated donors not only strongly increased anti-SARS-CoV-2 Ab titres and sero-neutralisation ratios but also increased cross-reactivity with a broader spectrum with respect to variants to which the donor has not been exposed.(12,29) High titer plasma from such convalescent vaccinated donors may be endowed with increased clinical efficacy. Early intervention with CCP has been associated with improved outcome.(3,4) Patients in our study exhibited a median of 7 days of symptoms at the time of inclusion, which is a short time period compared to most reported trials involving hospitalised patients. However, pre-specified subgroup analysis did not favour increased CCP efficacy associated with a shorter time period since symptoms onset. The high frequency of patients with underlying immunosuppression, for whom seroconversion is not expected early on, may contribute. Also, and as observed in other COVID-19 trials, early hospitalisation may be associated with more severe disease.(8,22)

Patients in the CCP arm tended to exhibit worsening pulmonary clinical conditions compared patients in the UC arm early after transfusion. The occurrence of early transient pulmonary worsening after CCP transfusion has been reported elsewhere as well (30), and may be in relation to an antibody-dependent enhancement involving immune-complex mediated inflammatory immunopathology in infected tissues.(31) Also, an Ab-dependent FcR-mediated infection of tissues macrophages (and circulating monocytes) may result in a massive inflammatory response, as recently evidenced (32), and may contribute as well. In fact, such early outcomes are seldomly reported in clinical studies. Furthermore, an early pulmonary worsening may be challenging to distinguish from transfusion associated circulatory overload (TACO), transfusion-related acute lung injury (TRALI) or overall disease worsening, possibly initiated before transfusion. The transfusion of 4 units of plasma may have contributed to circulatory overload in some patients. Further spacing of CCP administration (i.e. 1 unit / day over 4 days) could reduce such a risk. Importantly such early worsening did not prevent subsequent improvement and increased survival as early as day 14 post randomization, although not statistically significant. Of note, the observation that antibody-mediated SARS-CoV-2 uptake by monocytes/macrophages triggers inflammatory cell death and inhibition of viral replication may provide a mechanism for subsequent disease improvement.(33)

Our study has some limitations. The relatively small size of the trial limited the ability to appropriately assess outcomes such as patient mortality. Nevertheless, this did not prevent several important findings, notably regarding CCP treatment in immunosuppressed patients. Also, information regarding patient serostatus at inclusion was often unavailable, preventing meaningful findings in this regard.

Lastly, although the mean Ab ratio in transfused CCP in our study was well above the FDA threshold defining high titre CCP (Euroimmun anti-SARS-CoV-2 IgG ratio > 3.5) (34), transfusion of higher titre CCP, notably from convalescent vaccinated donors, may result in enhanced efficacy.(3,12,23,26)

In addition, the recent emergence of the omicron variant with its BA.1 to BA.5 subvariants has highlighted the risks associated with immune-resistant SARS-CoV-2 and loss of efficacy of available monoclonal antibodies.(11) While several months are necessary to produce one or more new monoclonal antibodies more suited to the evolution of circulating viral strains, convalescent plasma, notably from vaccinated donors, has demonstrated increased resilience to immune-resistant SARS-CoV-2 variants,(12,30) increased scalability as it may rely on existing collection infrastructure, as well as increased adaptability. The time between the onset of a COVID-19 variant and the availability of convalescent plasma from donors infected with the given variant disease is approximately four weeks.

These results, along with the recent data obtained from other trials and cohort studies may support further evaluation and consequent use of convalescent plasma in immunocompromised patients for whom therapeutic options are currently scarce. Accordingly, recent AABB guidelines suggest CCP transfusion in addition to standard of care for hospitalized patients with COVID-19 and pre-existing immunosuppression.(35)

## Data Availability

The data for this article will be made available after publication on request from any qualified researchers or academicians. The data include: analysed deidentified participant data, data dictionary, study protocol, statistical analysis plan, and the informed consent form, among other data. The data will be shared for 2 years after publication upon receipt of a request sent to.

## Acknowledgements

The authors wish to thank all physicians, nurses, and assistant nurses who took care of the patients, clinical research assistants and clinical research doctors who included and followed the patients during the trial, physicians, nurses and staff involved in convalescent plasma collection, manufacturing, testing and issuing, and above all the patients who agreed to participate in the study and the convalescent donors who generously gave their plasma. Special thanks to the International Direction of Clinical Research (DRCI) of Assistance Publique-Hôpitaux de Paris (APHP), the trial sponsor, and the Unité de Recherche de l’Est Parisien (URC-EST, APHP.SU, site St Antoine site), who managed the trial.

## Contributorship statement

KL and PT were involved in the protocol design and study design, including conceptualization, methodology, funding acquisition, and resources. NS and TS did the data curation, investigation, project administration, supervision, and validation. RP and GB did the data analysis. TH, AM, TC, SGL,FA, JS, GMB, JMM, FB, CC, OH, VP, MM, XL, PB, FP, DS, SR, LR and JMM included the patients. OH, XM, PLT, MRR and PR designed the CORIMUNO19 platform trial and conceptualized the embedded trials. The manuscript was drafted by KL and PT and read and edited by all authors who approved the final manuscript. RP and GB had direct access to data.

## ethics approval and consent statement

Ethical clearance was obtained from CPP Ile de France VI on April 10, 2020 (no. 26-20 Med.1°) and all patients gave their informed and signed consent to the inclusion and participation to the study.

